# Effect of Systemic Inflammatory Response to SARS-CoV-2 on Lopinavir and Hydroxychloroquine Plasma Concentrations

**DOI:** 10.1101/2020.07.05.20146878

**Authors:** Catia Marzolini, Felix Stader, Marcel Stoeckle, Fabian Franzeck, Adrian Egli, Stefano Bassetti, Alexa Hollinger, Michael Osthoff, Maja Weisser, Caroline E. Gebhard, Veronika Baettig, Julia Geenen, Nina Khanna, Sarah Tschudin-Sutter, Daniel Mueller, Hans H. Hirsch, Manuel Battegay, Parham Sendi

## Abstract

**Background:** Coronavirus disease 2019 (COVID-19) leads to inflammatory cytokine release, which can downregulate the expression of metabolizing enzymes. This cascade affects drug concentrations in the plasma. We investigated the association between lopinavir (LPV) and hydroxychloroquine (HCQ) plasma concentrations and the values of acute phase inflammation marker C-reactive protein (CRP).

**Methods:** LPV plasma concentrations were prospectively collected in 92 patients hospitalized at our institution. Lopinavir/ritonavir was administered 12-hourly, 800/200 mg on day 1, and 400/100 mg on day 2 until day 5 or 7. HCQ was given at 800 mg, followed by 400 mg after 6, 24 and 48 hours. Hematological, liver, kidney, and inflammation laboratory values were analyzed on the day of drug level determination.

**Results:** The median age of study participants was 59 (range 24–85) years, and 71% were male. The median duration from symptom onset to hospitalization and treatment initiation was 7 days (IQR 4–10) and 8 days (IQR 5–10), respectively. The median LPV trough concentration on day 3 of treatment was 26.5 μg/mL (IQR 18.9–31.5). LPV plasma concentrations positively correlated with CRP values (r=0.37, p<0.001), and were significantly lower when tocilizumab was preadministrated. No correlation was found between HCQ concentrations and CRP values.

**Conclusions:** High LPV plasma concentrations were observed in COVID-19 patients. The ratio of calculated unbound drug fraction to published SARS-CoV-2 EC50 values indicated insufficient LPV concentrations in the lung. CRP values significantly correlated with LPV but not HCQ plasma concentrations, implying inhibition of cytochrome P450 3A4 (CYP3A4) metabolism by inflammation.

## Introduction

Clinical trials have been launched to find effective treatment against the novel coronavirus, severe acute respiratory syndrome coronavirus 2 (SARS-CoV-2), the cause of the respiratory illness termed coronavirus disease 2019 (COVID-19) (1, 2). Approximately 15% of COVID-19 patients develop severe pneumonia (3). Cytokine release syndrome is an important factor for disease progression. Thus, treatment rationales for COVID-19 have focused on both antiviral activity and control of the infection-induced cytokine storm (4). Direct interaction between the 2 modalities must be evaluated, however, because infectious and inflammatory diseases have an impact on drug metabolism (5, 6).

The release of inflammatory cytokines such as interleukin-6 (IL-6) activates intracellular signaling cascades, leading to the downregulation of cytochrome P450 enzymes (CYPs) (7). The decrease in expression and activity of CYPs is explained by transcriptional suppression of CYP mRNA, triggering a decrease in enzyme synthesis (5, 6). Systemic inflammation affects CYPs differently with a more pronounced decrease in CYP3A4 expression followed by CYP2B6, CYP2C19, CYP2C9, CYP2D6 and CYP1A2 (5, 6, 8). Correlations have been reported between elevated C-reactive protein (CRP) values and high plasma levels of antipsychotic drugs (9) and voriconazole (10). CRP production is triggered by IL-6, and conversely, IL-6 suppression can be monitored with plasma CRP levels (11).

The HIV drug lopinavir/ritonavir (LPV/r) has been repurposed for the treatment of SARS-CoV-2 (2). Recent brief reports of 8 (12), 12 (13), and 21 (14) COVID-19 patients, noted considerably higher LPV plasma concentrations than those observed in HIV patients (15). Considering the inhibition of drug metabolism by cytokine release, and the administration of LPV/r (metabolized by CYP3A4), we had the rationale to prospectively monitor LPV plasma concentrations in our cohort of COVID-19 patients.

The objective of this study was to investigate the association between CRP values and LPV plasma concentrations in COVID-19 patients. With this approach, we aimed to underscore the hypothesis that high inflammatory markers in the blood correlate with high LVP plasma concentrations. For comparison, we measured hydroxychloroquine (HCQ) concentrations, because it is characterized by a different metabolism (16). We also performed an age-stratified analysis to explore the combined effect of aging and inflammation on drug plasma levels. Finally, we discuss our LPV plasma trough concentration results in the context of calculated unbound concentrations in the lung compartment and published 50% effective concentrations (EC50) values for SARS-CoV-2.

## Methods

All adults (≥18 years) hospitalized at the University Hospital in Basel between 25 February and 30 April 2020 for a COVID-19 infection (confirmed by real-time reverse transcriptase-polymerase chain reaction [RT-PCR] from nasopharyngeal swab specimens) were screened for study eligibility. The study was part of a COVID-19 cohort consortium investigation and approved by the northwest/central Switzerland Ethics Committee (EKNZ 2020-00769).

### Study population

COVID-19 patients were eligible, if they were treated with LPV/r. Patients were excluded if LPV drug concentrations were not measured. Prior to administration of LPV/r and HCQ, all concomitant drugs were reviewed for potential drug-drug interactions (DDI) via a website incorporated in our institutional treatment recommendations (17). Concomitant intake of CYP3A4 inhibitors or inducers was stopped or switched to another compound with similar therapeutic effect. Corticosteroids were not administered, except in 3 individuals in whom a low-dose long-term treatment with prednisone was continued (5 mg/d in 2 patients, 10 mg/d in 1 patient). Other drugs affecting inflammation were not administered, with the exception of tocilizumab (TCZ). *Treatment concepts for COVID-19 and dosing rationale*: Our institutional treatment recommendations include the administration of LPV/r and HCQ for hospitalized patients. To achieve rapidly high LPV/r plasma concentrations, we administered a double dose in the first 24 hours. This approach in the early treatment phase was presumed necessary to suppress the high SARS-CoV-2 viral load in the early stage of disease (“hit early and hit hard”). The LVP/r treatment schedule included 800/200 mg twice daily on day 1, followed by a maintenance dose of 400/100 mg every 12 hours for another 4 to 6 days. LVP/r treatment was combined with HCQ for 2 days (i.e., 800 mg loading dose followed by 400 mg at 6 hours, 24 hours, and 48 hours). In patients with clinical signs and findings suggestive for COVID-19-induced hyper-inflammation, the use of TCZ was considered at the discretion of the treating COVID-19 care team. Parameters for consideration were defined in the institutional diagnostic recommendations for COVID-19. They included clinical signs (breathing frequency ≥30 per minute, O2 saturation <93%), laboratory results (CRP ≥75mg/L) and the extent of radiological findings in the computed tomography scan of the lung (typical ground-glass opacities, infiltrates in ≥ 4 lobes or considerable progression of infiltrates within 24 to 48 hours). TCZ was administered intravenously at a dose of 8 mg/kg body weight, with single dose or 2 doses within 24 hours.

### Quantification of LPV and HCQ plasma concentrations

The institutional diagnostic recommendations for COVID-19 suggest obtaining LVP plasma trough levels on day 2 or 3 of treatment. LPV levels were quantified by using commercial calibrators and controls for liquid chromatography mass spectrometry methods (Recipe Chemicals + Instruments, Munich, Germany). The lower limit of quantification was 0.1 μg/mL.

HCQ levels were measured from available plasma material obtained for LPV trough determination. HCQ was quantified with a validated liquid chromatography mass spectrometry method developed by the laboratory of clinical chemistry at the University Hospital in Zurich, Switzerland. The lower limit of quantification was 10 ng/mL.

### Data management, variable categorization, and statistical analysis

Patient demographics, laboratory data, vital parameters, and medication records were extracted from the electronic medical reports and the institutional Clinical Data Warehouse.

Information on the time interval between onset of symptoms consistent with COVID-19 and (i) hospitalization and (ii) initiation of antiviral treatment were investigated prospectively. Laboratory results obtained at the day of drug level measurement were used for this analysis.

Because age-related physiological changes can affect drug pharmacokinetics (18), we categorized patients as <65 years or ≥ 65 years. As indicated earlier, we used a tentative CRP cutoff value of 75 mg/L to aid decision making for the administration of TCZ. This CRP level was used as marker for inflammation for the analysis in the study (i.e., <75 vs ≥ 75 mg/L).

In patients receiving TCZ prior to the measurement of LPV or HCQ plasma concentrations, a time interval cutoff of 12 hours for inflammation inhibition and consecutive effect on drug metabolism was predefined. This value was chosen after consideration of various parameters (i.e., presumed time to clinical resolution of cytokine release syndrome after TCZ administration (19), dynamics of CRP levels in infections (20), drug administration schedule). Hence, in the case of TCZ administration at ≤12 hours prior to the measurement of LPV trough levels, the interval between the two time points was considered to be too short for having an effect on LPV plasma concentrations.

Absolute numbers, percentages, medians, and interquartile ranges (IQRs) were used to report demographic characteristics and laboratory results. The Mann-Whitney U test was used to compare continuous data, and the Spearman correlation coefficient to explore associations of interest. All statistical analyses were performed with GraphPad Prism and SPSS.

## Results

Of 170 COVID-19 patients hospitalized in our institution within the study time frame, 92 RT-PCR confirmed positive cases with available LPV plasma concentrations were included in the study. The median age of study participants was 59 (IQR 48–70; range 24– 85) years, and the majority were males (71%). The median time from onset of symptoms to hospitalization was 7 (IQR 4–10) days, and from onset of symptoms to initiation of LPV/r and HCQ treatment, was 8 (IQR 5–10) days. Twenty-seven (29%) individuals were transferred to the intensive care unit (ICU) during the hospitalization. Overall, 35 (38%) patients received TCZ, 19 (54%) prior to LPV plasma concentration measurement and 16 (46%) afterward. The median CRP values at the day of LPV plasma measurements in these TCZ groups were 88.9 (IQR 48.2-153.2) mg/L, 79.9 (IQR 48.2-129.6) mg/L and 105.4 (IQR 51.9-153.7) mg/L, respectively. In 3 individuals who received TCZ before measurement of LPV plasma concentrations, the time interval between the two time points was ≤12 hours. For analysis purposes, the LPV plasma levels of these 3 patients were assigned to the group who received TCZ after drug level measurement. The CRP values in these individuals were 44.2, 124,8 and 165.8 mg/L.

Patients admitted to the ICU tended to have a higher body weight, lower albumin and hemoglobin levels, higher creatine kinase and CRP values than did patients who were not treated in the ICU (**Table 1**). Twenty (22%) patients presented with moderate or severe renal impairment. **Table 1** shows the demographic and clinical characteristics of the patients.

**Table 1.**
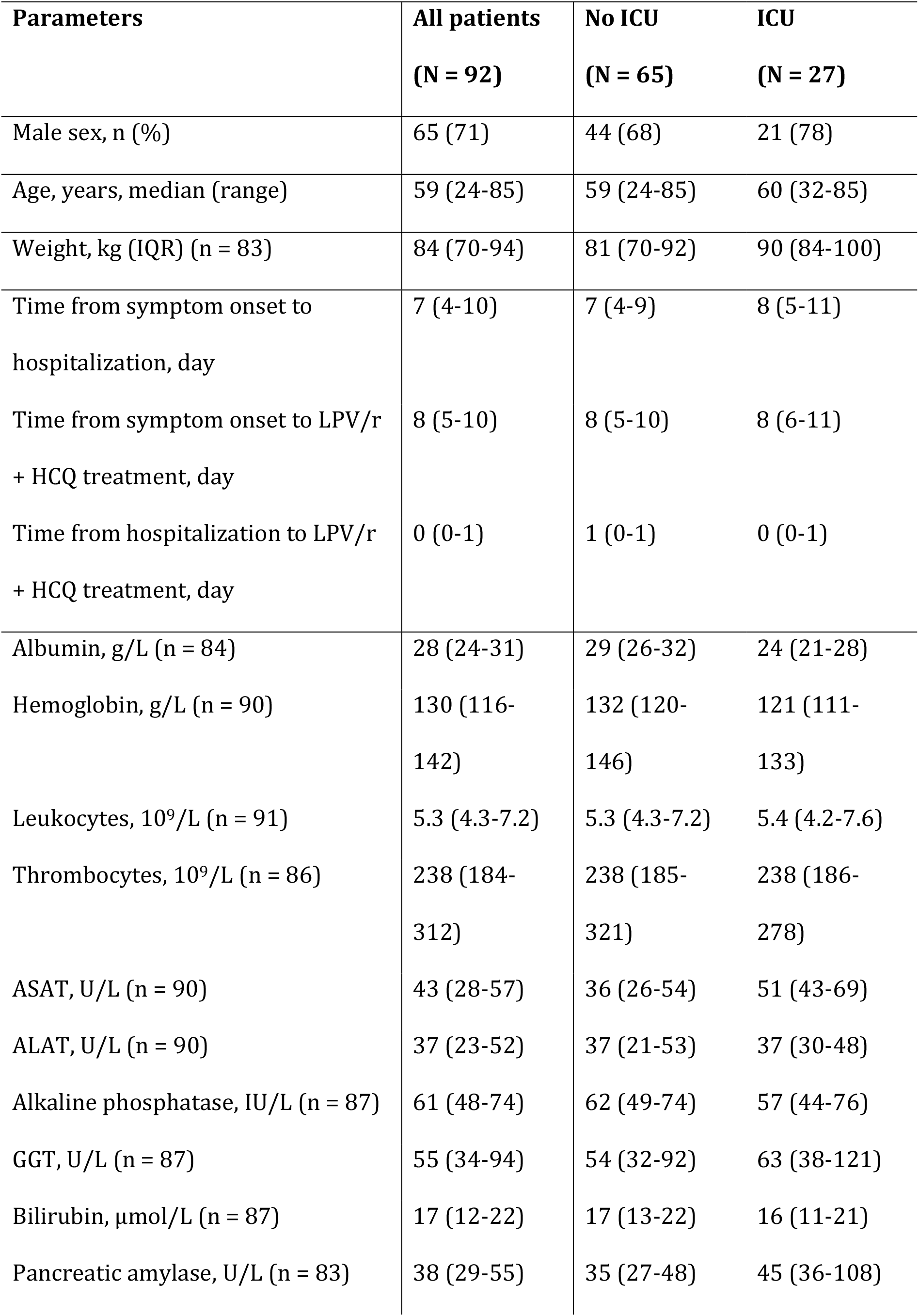
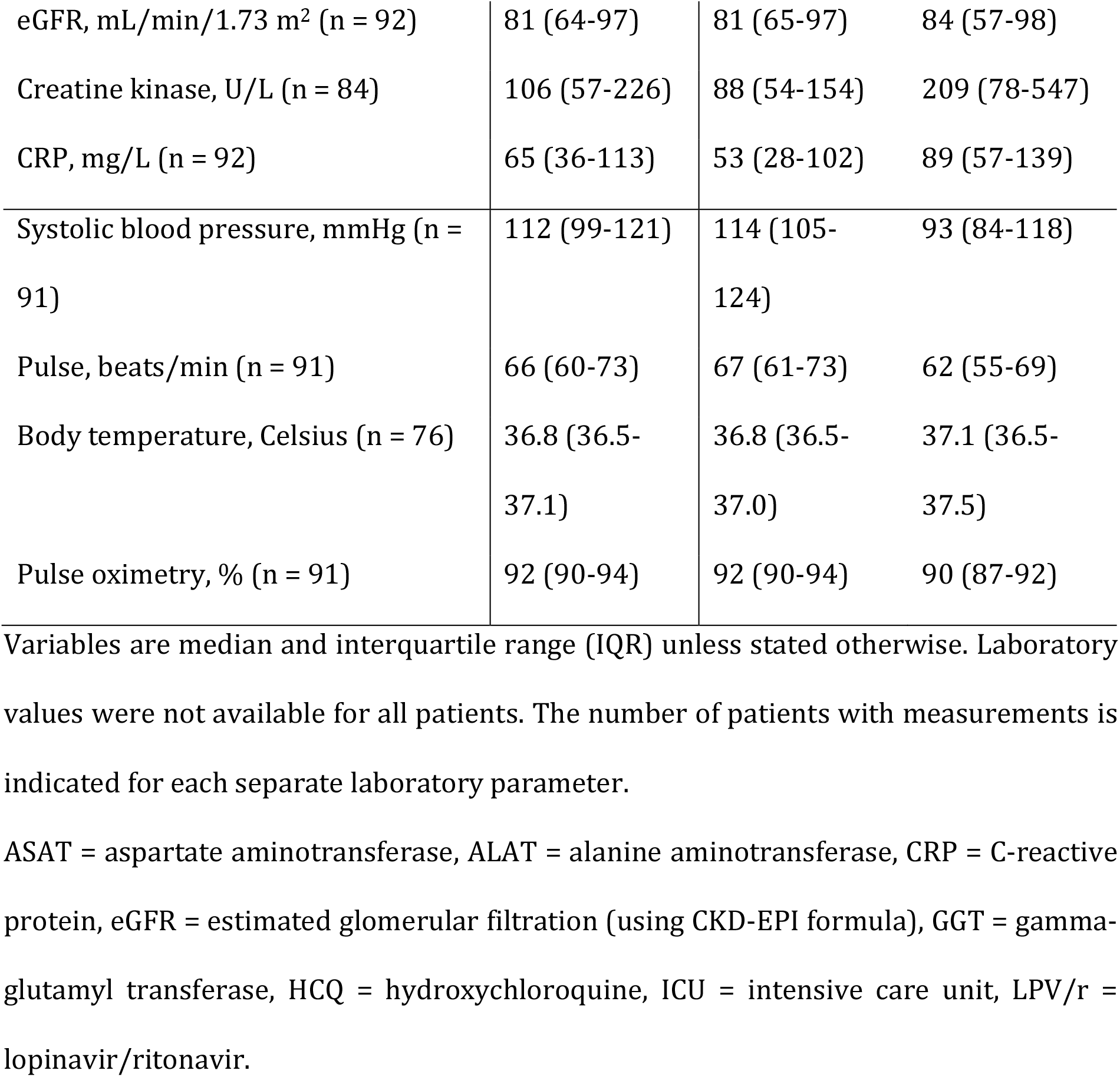
Demographic, clinical and laboratory characteristics of the study population on the day of LPV plasma concentration measurement.

### LPV levels and impact of inflammation

LPV trough levels (12 h ± 3 hours after the last drug intake) ranged from 7.7 to 42.3 μg/mL with a median value of 26.5 (IQR 18.9–31.5) μg/L (**Figure 1**). LPV plasma concentrations were measured after a median time of 3 (IQR 3– 4) days and correlated positively with CRP values (r=0.37, p<0.001, 92 observations) and leukocytes (r=0.32, p=0.002, 91 observations). When stratifying patients by predefined CRP level, we observed significantly higher LPV concentrations in patients with CRP ≥75 mg/L than in those with <75 mg/L (median levels: 30.7 vs 20.9 μg/mL, p < 0.001) (**Figure 2A**). TCZ administration >12 hours prior to LPV measurement demonstrated significantly lower LPV plasma concentrations (median 18.7 μg/mL) than did the comparison group (i.e., no TCZ administration or TCZ administration ≤12 hours prior to LPV measurement) (median 28.8 μg/mL, p<0.001, **Figure 3**). No other significant correlations were found with any other parameters listed in **Table 1**.

**Figure 1.**
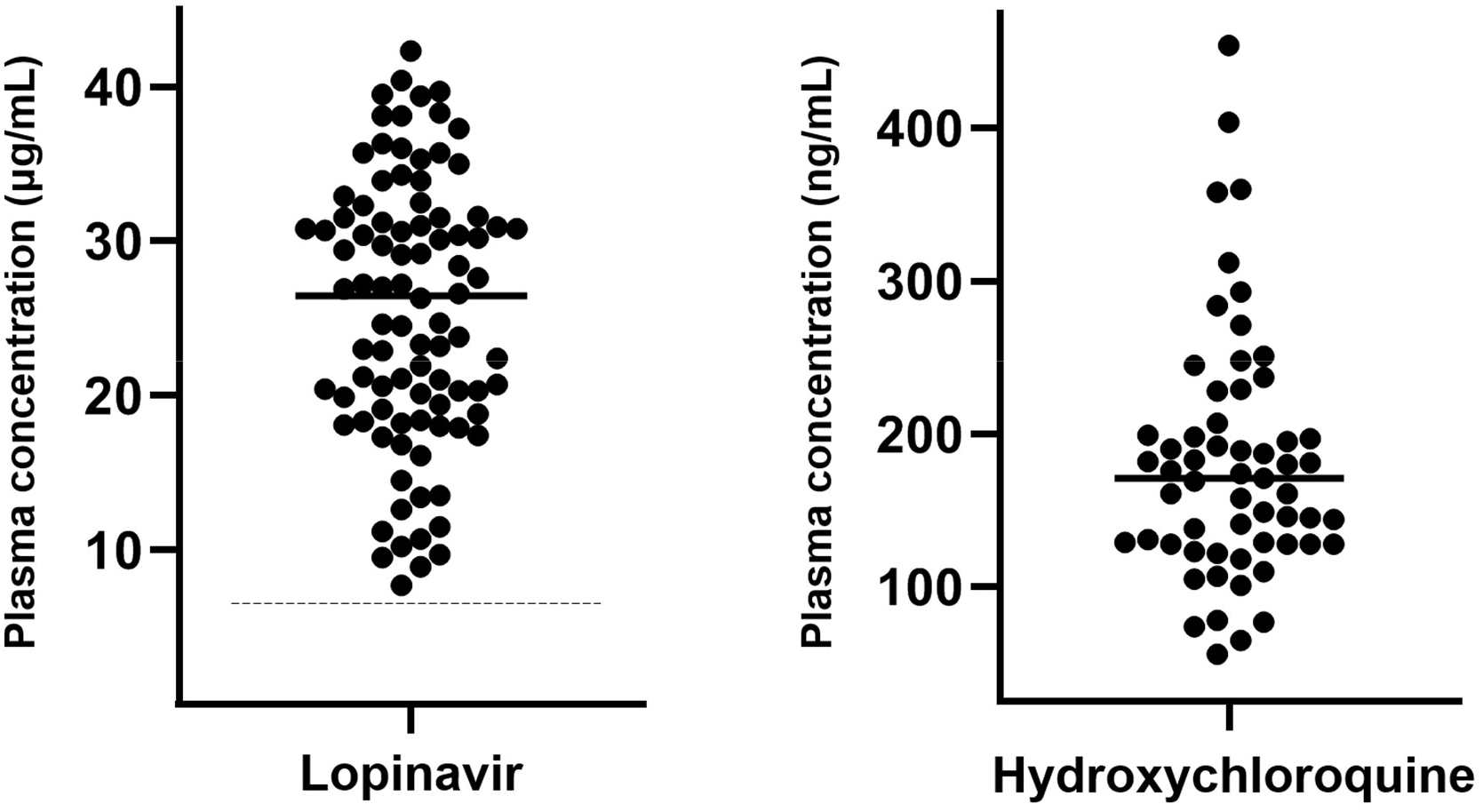
Lopinavir (n = 92) and hydroxychloroquine (n = 59) plasma concentrations in COVID-19 patients. The medial Lopinavir plasma concentration was 26.5 (IQR 18.9–31.5) μg/mL. The median Hydroxychloroquine plasma concentration was 171 (IQR, 128–207) ng/mL. The dashed line represents the historical lopinavir trough level observed in HIV-infected individuals treated with lopinavir/ritonavir 400/100 mg twice daily (i.e., 7.1 μg/mL) (15).

**Figure 2.**
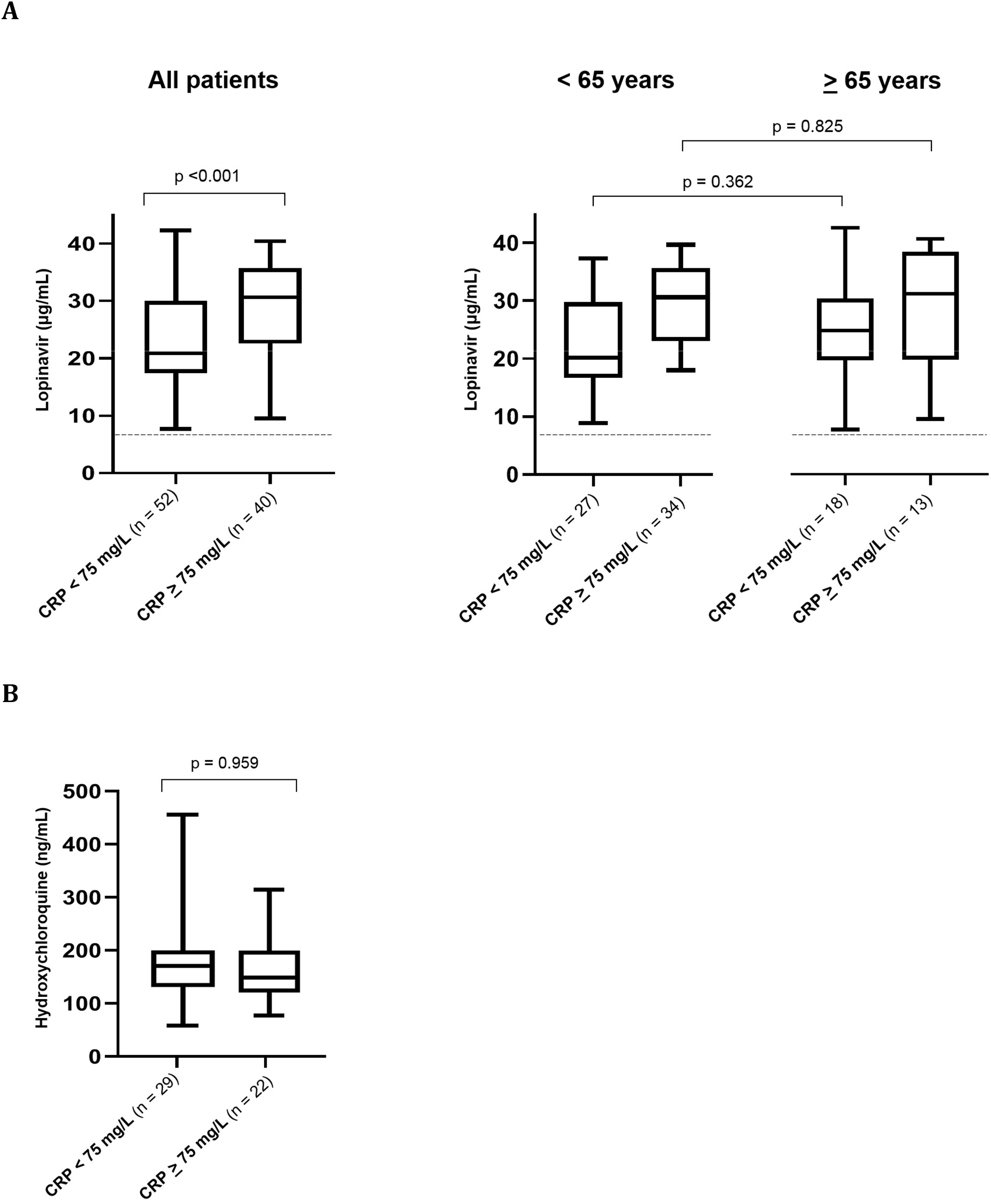
Box plots (showing the 5^th^, 25^th^, 50^th^ 75^th^ and 95^th^ percentiles) of lopinavir trough concentrations by CRP values in all patients and by age group (A) and box plots of hydroxychloroquine concentrations by CRP values for COVID-19 patients with trough levels (B). CRP = C-reactive protein. The dashed line represents the historical lopinavir trough level observed in HIV-infected individuals treated with lopinavir/ritonavir 400/100 mg twice daily (i.e., 7.1 μg/mL) (15).

**Figure 3.**
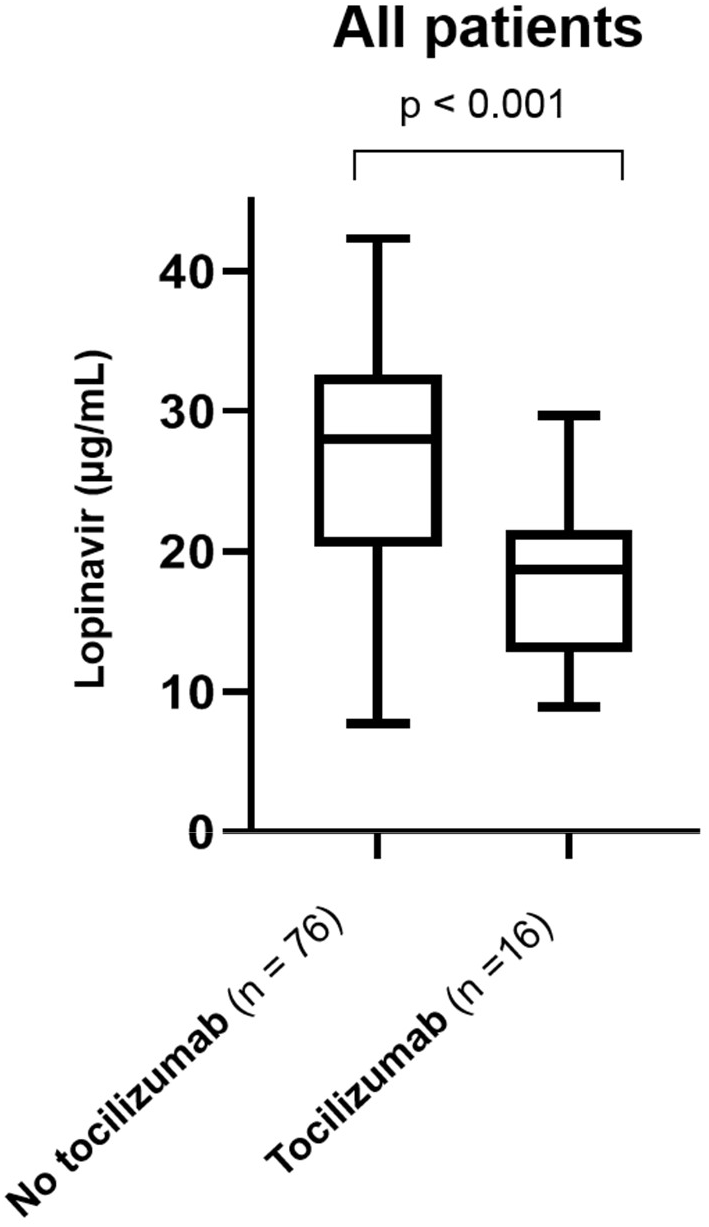
Box plots (showing the 5^th^, 25^th^, 50^th^ 75^th^ and 95^th^ percentiles) of lopinavir plasma trough concentrations in COVID-19 patients by administration of tocilizumab. The left bar includes LPV plasma levels from COVID-19 patients with no TCZ administration (n = 57) or TCZ administration < 12 hours prior to LPV measurement (n = 19), (median 28.8 µg/mL). The right bar represents LPV samples from COVID-19 patients with TCZ administration > 12 hours prior to LPV measurement (median 18.7 µg/mL).

### Combined effect of age and inflammation on LPV concentrations

Median LPV plasma trough levels were insignificantly higher in patients who were ≥65 years (26.9 μg/m, n = 33) than in those who were <65 years (24.5 μg/mL, n = 59) (**Figure 2A**). Accordingly, median LPV concentrations were not different in patients with CRP values ≥75 mg/L and who were ≥65 vs <65 years (median levels: 31.0 vs 30.6 μg/mL, p = 0.825) or in patients with CRP values <75 mg/L who were ≥65 vs <65 years (median levels: 24.7 vs 20.2 μg/mL, p=0.362) (**Figure 2A**).

### HCQ concentrations

HCQ concentrations were measured in 59 patients from available plasma samples, and ranged from 56 to 454 ng/mL with a median value of 171 (IQR 128– 207) ng/mL (**Figure 1**). In 51 plasma samples, the median time interval since the last drug intake was of 22 (range 12–31, IQR 14–23) hours, and the values showed no correlation with CRP values (r=0.044, p=0.76) or any other laboratory parameter listed in **Table 1**. HCQ plasma concentrations were not statistically different in patients with CRP values <75 or ≥75 mg/L (median levels: 149 versus 148 ng/mL, p=0.959) (**Figure 2B**). There was no correlation between LPV and HCQ plasma concentrations (r=0.197, p=0.166, n=51).

## Discussion

Median LPV trough concentrations were an unexpected 3.5-fold higher in patients infected with SARS-CoV-2 than in HIV-infected patients, as reported historically (i.e., 7.1 μg/mL) (15). Our prospective analysis on LPV plasma concentrations in 92 patients is in line with recent observations in small series that reported LPV plasma concentrations from 13 to 18 μg/mL in COVID-19 patients (12-14). The even higher trough concentrations in our study (i.e., median 26.5 μg/mL) might be explained by the double dose LPV/r dose (800/200 mg) at day 1 and the differences in the severity of COVID-19 between the studies. The median CRP values available in two of the aforementioned brief reports were 13.6 mg/L (12) and 48.9 mg/L (13) vs 65 mg/L in our study.

We investigated possible reasons for high LPV plasma concentrations in COVID-19 patients. HCQ-mediated inhibition of the hepatic organic anion transporting polypeptide 1A2 (OATP1A2) (21), and interference with liver entry and subsequent metabolic elimination is, in our view, not plausible. OATP1A2 is expressed on the apical membrane of cholangiocytes, where it reabsorbs drugs excreted into the bile (22). Inhibition of this transporter would likely facilitate LPV/r biliary elimination. Viral-induced liver damage may cause impaired drug metabolism and high LPV plasma concentrations. However, the vast majority of individuals in our study population had only mildly elevated liver enzymes (**Table 1**). The effect of the double dose within the first 24 hours on LPV plasma trough concentrations measured after a median time of 3 (IQR 3-4) days is difficult to assess. LPV/r 800/200 mg single dose pharmacokinetic studies reported concentrations <12 μg/mL (23) or <14 μg/mL (24), 12 hours after intake. In a study with healthy HIV-negative volunteers, LPV trough levels ranged from 8.3 to 13.8 μg/mL at day 2 of treatment with 800/200 mg twice daily (25). In our study population, 81 (88%) samples had LPV plasma levels >14 μg/mL, 66 (72%) >20 μg/mL and 35 (38%) > 30 μg/mL. these data together with the LPV pharmacokinetics data in the literature (23-25), noticeably suggests that the elevated LPV trough concentrations observed in COVID-19 patients cannot be explained only by the effect of the initial double dose. Our findings support the hypothesis that the systemic inflammatory response in COVID-19 patients inhibits drug metabolism, leading to elevated LPV plasma concentrations. Conversely, blocking inflammation with TCZ was associated with lower LPV plasma concentrations. This is possibly explained by the fact that TCZ inhibition of inflammatory cytokines leads to a normalization of CYP metabolism.

Aging is associated with physiological changes and decline of the immune function, which altogether can impact drug pharmacokinetics (18). However, LPV plasma trough concentrations were not significantly different in patients who ≥ 65 than in those who were <65 years in our study.

Inflammation has been shown to have the greatest impact on CYP3A4 expression (7). This increase may, in turn, impact the magnitude of DDIs, because LPV/r inhibits CYP3A4 in a concentration-dependent manner (26). Co-administered CYP3A4 substrates can – *per se* – be also affected by inflammation, and can further increase the magnitude of DDIs. This interaction is illustrated by a case series of 12 patients who were followed up for direct oral anticoagulant treatment (DOAC) before and after being infected with SARS-CoV-2. LPV/r was started while DOAC was maintained at the same dose. DOAC levels after initiation of LPV/r treatment showed an average 6-fold increase (27). The co-administration of the strong CYP3A4 inhibitor ritonavir has been shown to increase rivaroxaban levels by 2.5-fold in healthy volunteers (28), whereas rivaroxaban plasma concentrations were increased by 7-up to 31-fold in COVID-19 patients treated with LPV/r (27). Notably, the disappearance of the inhibitory effect on CYP3A4 may take up to 5 days after stopping LPV/r (29).

The high LPV plasma concentrations observed in COVID-19 patients inevitably raise the question about the LPV levels that can be achieved in the lung. LPV/r is thought to act by inhibiting the enzyme 3-chymotrypsin-like protease (3CL^pro^) of SARS-CoV-2, thereby disrupting the cleavage of the viral protein and release from the host cell (30). Coronavirus proteases, including 3CL^pro^, do not contain a C2-symmetric pocket, resulting in an unspecific inhibition (31). Recently, Choy et al. (32) investigated the EC50 of LPV in inhibiting SARS-CoV-2 replication in Vero E6 cells. The cells were treated with the compound for 1 hour prior to the infection by the virus at a multiplicity of infection of 0.02. The authors reported EC50 values of 26.63 and 26.10 μM, measuring infectious virus and viral RNA, respectively. These values correspond to *in-vitro* concentrations of 16.7 and 16.4 μg/mL respectively (32). The antiviral activity *in vivo* is estimated by calculating the ratio of unbound drug concentrations achieved in the lung at the administered dose to the *in vitro* EC50 value (R_LTEC_) (33). LPV plasma measurements in 12 COVID-19 patients showed median total and unbound trough concentrations of 18.0 μg/mL and 0.16 μg/mL, respectively, resulting in an unbound fraction of 0.88% (13). This fraction is consistent with the results from a previous study (34). The simultaneous determination of LPV in epithelial lining fluid (ELF) and plasma indicated an ELF/plasma ratio of 1.77 (35). Considering our total observed LPV trough plasma concentrations, the extrapolated unbound LPV trough level is 0.23 μg/mL. This value corresponds to an unbound LPV level in ELF of 0.41 μg/mL, which gives a R_LTEC_ of 0.025. Even though the majority of the observed total LPV plasma concentrations in COVID-19 patients were above the published EC50 values for SARS-CoV-2 (32), boosted LPV is unlikely to attain sufficient effective levels in the lung to inhibit the virus. In line with these arguments, current available clinical data do not demonstrate evidence for the efficacy of LPV/r for COVID-19 (36, 37). HCQ has been used historically for malaria and immune diseases. Its ability to inhibit SARS-CoV-2 is thought to be due to an increase in endosomal pH, thereby impairing the entry of the virus into the cell. HCQ also interferes with the glycosylation of cellular receptors for SARS-CoV-2, resulting in reduced virus-cell binding. Finally, HCQ has immunomodulatory activity that may suppress the cytokine storm (16).

The median HCQ concentrations observed in our study (i.e., 171 ng/mL, IQR 128–207) is comparable to those reported in another study with COVID-19 patients (220 ± 110 ng/mL) (38), and to steady-state trough levels observed in patients with lupus erythematosus (i.e., 103–130 ng/mL)(39). Thus, the HCQ plasma concentrations in COVID-19 patients, in contrast to reported LPV plasma concentrations, were not higher than those previously observed in studies with other indications. Furthermore, no correlation was observed with CRP values. This difference may possibly be explained by the different metabolic pathways of HCQ and LPV/r, as inflammation affects CYPs differently (7). Furthermore, HCQ is known to have higher concentrations in tissue than in plasma (approximately 200-to 700-fold higher), resulting in a large distribution volume and a long half-life (33). Therefore, HCQ plasma concentrations from COVID-19 patients might not be suitable to reflect the effect of inflammation given that HCQ does not achieve steady-state concentrations during the short treatment course. Similar to LPV/r, HCQ was shown to have a low R_LTEC_ (i.e., 0.11–0.34), indicating that HCQ levels achieved *in vivo* do not result in adequate clinical activity against SARS-CoV-2 (33). These calculations are supported by a studies failing to demonstrate a benefit of HCQ in both hospitalized patients with COVID-19 (40), and as prophylaxis after SARS-CoV-2 exposure (41).

Some limitations of this study should be acknowledged. We did not consider IL-6 measurement as a routine diagnostic value within our COVID-19 cohort, and hence, in study. IL-6 is a central mediator of the acute-phase response and a primary determinant of hepatic production of CRP. IL-6 has many other pathophysiologic roles in humans (42) and its diagnostic value for COVID-19, in particular for non-severe cases, is unknown. The selection of cutoff of 12 hours in the case of TCZ administration prior to measurement of LPV plasma concentrations was clinically reasonable but arbitrary. However, this limitation applied to only 3 patients, and had no statistical influence on the results.

In conclusion, high LPV trough plasma concentrations were observed in COVID-19 patients. However, the calculated unbound concentrations in the lung indicates insufficient levels to inhibit SARS-CoV-2 replication. LPV levels correlated positively with CRP values and negatively with the preadministration of TCZ, indicating that COVID-19 related cytokine release significantly inhibits CYP3A4. Caution is advised when prescribing CYP3A4 substrates with a narrow therapeutic index to COVID-19 patients because of the risk of elevated drug levels and related toxicities.

## Data Availability

Data referred to in the manuscript are available

## Acknowledgements

We thank all the healthcare professionals and personnel in our institution who are involved in organizational processes and patient care during the COVID-19 pandemic. We are indebted to Prof. Dr. sc. nat Katharina Rentsch, Laboratory Medicine, University Hospital Basel, for her valuable role in acquiring plasma drug levels.

